# Prediction of Obstructive Sleep Apnea Using Variational Autoencoder-Derived Features From 12-Lead ECG

**DOI:** 10.64898/2026.09.22.26363710

**Authors:** Christopher Harvey, Amulya Gupta, Barsha Halder, Jeffrey Thompson, Soniya Mishra, Matthew KP Gratton, Damien Stevens, Amit Noheria, Diego R. Mazzotti

**Affiliations:** Program for AI & Research in Cardiovascular Medicine (PARC), Department of Cardiovascular Medicine, University of Kansas Medical Center, Kansas City, KS; Department of Biostatistics and Data Science, University of Kansas Medical Center, Kansas City, KS; Division of Medical Informatics, Department of Internal Medicine, University of Kansas Medical Center, Kansas City, KS; Division of Pulmonary, Critical Care and Sleep Medicine, Department of Internal Medicine, University of Kansas Medical Center, Kansas City, KS; University of Kansas, Lawrence, KS

**Keywords:** obstructive sleep apnea, electrocardiogram, machine learning, gradient boosting, variational autoencoder, representation learning

## Abstract

**Background:** Obstructive sleep apnea (OSA) is a highly prevalent sleep disorder. Major risk factors include obesity, older age, and male sex. Given the well-established association between OSA and cardiovascular disease, markers of cardiovascular health derived from electrocardiography (ECG) may help identify individuals with undiagnosed OSA. This study evaluated whether 12-lead ECG features improve machine learning-based prediction of OSA beyond established clinical factors.

**Methods:** We conducted a retrospective cohort study of individuals who underwent diagnostic sleep studies at the University of Kansas Health System Sleep Disorders Center (2004-2025) and had a 12-lead ECG acquired within one year. Standard ECG features (intervals, amplitudes, voltage-time integrals, morphology descriptors) were extracted from the representative beat. Latent representations were derived using two variational autoencoder (VAE) models pretrained on >1 million ECGs: a 30-dimensional representative-beat VAE and a 90-dimensional full 10-second ECG VAE. Three LightGBM models were trained: clinical factors only (age, sex, weight, height, body mass index [BMI], Epworth sleepiness scale [ESS]), ECG features only, and a combined model. Feature selection and hyperparameter tuning were performed separately for each model within a 5-fold cross-validation scheme. The outcome was moderate-to-severe OSA (AHI≥15 events/h), evaluated by AUROC (bootstrapped 95% CI) in a 10% holdout test set.

**Results:** The dataset included 7,699 individuals (52.2% women, mean age 58.0 ± 14.2 years, BMI 33.6±8.2 kg/m², ESS 8.8±5.1; 36.4% with moderate-to-severe OSA) Training AUROC (95% CI) was 0.71 (0.66-0.76) for the combined model, versus 0.65 (0.59-0.70) for the clinical-only model and 0.66 (0.61-0.72) for the ECG-only model. Holdout test AUROC was 0.78 (combined), 0.69 (clinical-only), and 0.71 (ECG-only). Feature importance identified age, BMI, weight, ECG VAE latent variables, height, sex, and P-wave voltage-time integral as relevant contributor to the predictions.

**Conclusions:** Latent ECG features extracted by VAE improve prediction of moderate-to-severe OSA when combined with clinical factors.

## Introduction

Obstructive sleep apnea (OSA) is one of the most prevalent sleep disorders worldwide, affecting nearly 1 billion adults aged 30-69 years [1]. The condition arises from recurrent partial or complete collapse of the upper airway during sleep, causing oxygen desaturation, arousal from sleep, and sympathetic activation [2]. OSA is associated with a broad spectrum of cardiometabolic consequences including hypertension, coronary artery disease, cardiac arrhythmia, heart failure, and metabolic syndrome [3]. Proposed mechanisms include intermittent hypoxemia, sympathetic hyperactivation, endothelial dysfunction, oxidative stress, systemic inflammation, and the mechanical effects of repetitive negative intrathoracic pressure [4]. Randomized controlled trials of continuous positive airway pressure have not demonstrated reductions in major adverse cardiovascular events, possibly due to phenotypic heterogeneity and suboptimal adherence [5–7]. Nevertheless, identifying moderate-to-severe OSA carries clinical importance, as this group bears the greatest cardiovascular risk and is often the primary target for intervention [8,9].

Despite its high prevalence, most individuals with OSA remain undiagnosed. Current OSA screening relies on self-report questionnaires and anthropometric measures [10,11]. These tools offer only modest discriminative performance for moderate-to-severe disease [12] and capture no direct cardiovascular physiological information. ECGs are routinely acquired in outpatient cardiology and primary care settings, and their widespread availability makes them an attractive adjunct for OSA screening. ECG-derived features reflecting autonomic tone, ventricular hypertrophy, and conduction abnormalities [13] may encode information predictive of OSA beyond what is captured by demographic and anthropometric factors alone [14,15]. Prior studies have demonstrated that heart rate variability (HRV) parameters are associated with OSA and its severity [16,17], and deep learning models applied to overnight ECG recordings have shown promise for event-level apnea detection [18,19]. However, most prior approaches used sleep-acquired ECG signals rather than clinic-based 10-second recordings.

Moreover, the incremental value of conventional ECG morphology features and latent ECG representations over clinical predictors has not been systematically evaluated in large clinical cohorts.

Recent advances have demonstrated that ECG signals contain latent information predictive of structural and functional cardiovascular phenotypes, including left ventricular hypertrophy and systolic dysfunction, beyond conventional ECG measurements [20]. Variational autoencoder (VAE) models trained on large-scale ECG datasets can compress complex ECG waveforms into low-dimensional latent representations that preserve subtle morphological characteristics and enable highly effective downstream prediction tasks [21]. We therefore hypothesized that ECG-derived features, including VAE-based latent representations, may capture cardiovascular signatures of OSA and improve prediction of moderate-to-severe OSA beyond established clinical risk factors alone. To assess this hypothesis, we leveraged a single-center clinical cohort with paired diagnostic sleep studies and ambulatory 12-lead ECGs to test whether ECG-derived features improve prediction of moderate-to-severe OSA. We proposed that combined models would outperform clinical-only or ECG-only models. Finally, we examined feature importance to identify clinically interpretable ECG predictors of OSA.

## Methods

### Study participants

We conducted a retrospective study of individuals who underwent diagnostic sleep studies at The University of Kansas Health System (UKHS) Sleep Disorders Center between 2004 and 2025 and had at least one 12-lead ECG performed within one year of their sleep study. This study integrated two ongoing clinical data repositories at UKHS: one for diagnostic sleep studies and one for ECGs. Participants were identified based on the availability of both diagnostic tests during the study period. The study was approved by the University of Kansas Medical Center Institutional Review Board (STUDY00146506).

### OSA outcome definition

OSA was diagnosed based on in-laboratory polysomnography (SomnoStar SleepSystem, Viasys Healthcare, Conshohocken, Pennsylvania; Natus Sleepworks, Middleton, Wisconsin) or home sleep apnea testing (Nox-T3, Nox Medical Reykjavik, Iceland; WatchPAT, Itamar Medical Ltd., Caesarea, Israel) per AASM Scoring Manual guidelines [22]. Hypopneas required ≥30% reduction in airflow associated with ≥4% oxygen desaturation over at least 10 seconds. Participants were classified as no-to-mild OSA (AHI <15 events/h) or moderate-to-severe OSA (AHI ≥15 events/h), the binary outcome for all models. This threshold was selected because moderate-to-severe OSA carries the greatest cardiovascular risk burden [8] and is the primary target for intervention. Clinical covariates recorded at the time of sleep study included age, sex, weight, height, body mass index (BMI), and Epworth sleepiness scale (ESS) scores [23].

### ECG acquisition and signal processing

Clinically obtained standard 12-lead, 10-second ECG signals (500 Hz) and 1.2-second representative or median beats (1000 Hz) generated by the Philips® DXL (through July 2022) or GE® MUSE (August 2022 onwards) algorithms respectively were extracted for each patient. Orthogonal X, Y, and Z leads were reconstructed from the 12-lead ECG using the Kors matrix transformation [24]. A scaler 3D lead was computed as the Euclidean norm (L2 or root-sum-squares) of instantaneous voltages across X (right to left), Y (cranial to caudal), and Z (anterior to posterior) leads. A comprehensive set of ECG features (intervals and P-, QRS-, and T-wave specific amplitudes, voltage-time integrals, morphology descriptors) was extracted from representative beats across 16 leads (12 standard, 3 orthogonal leads, and 1 derived 3D lead).

### VAE models

Latent representations were derived using two VAE models pretrained with self-supervised learning for signal reconstruction on 1.18 million unselected Phillips ECGs from approximately 480,000 unique patients (2008-April 2022) to optimize reconstruction fidelity trained on the X, Y, and Z leads’ raw signals: (A) a representative-beat VAE producing 30 latent variables, and (B) a full 10-second ECG VAE producing 90 latent variables [21]. These latent features were incorporated alongside other extracted ECG features as inputs to the classification models.

The VAE models were trained in TensorFlow and consist of an encoder and a decoder. The encoder network comprised four 2D convolutional layers with filter sizes sequentially of 64, 64, 128, and 128. Each layer used a filter width of 15, a stride of 2, ELU activation function, and batch normalization. The final convolutional layer was flattened and passed to two fully connected layers with L2 regularization weighted 0.01 and ELU activations with dropout 0.1. The final layer was sampled using the population mean and log variance to obtain latent embeddings. The embeddings were then passed to the decoder made of 2 fully connected layers with L2 regularization, ELU activations and dropout, followed by four 2D transpose convolutional layers with filter sizes 128, 64, 32, 1, filter widths of 15, and strides of (1; 1; 1,2; 1,2) with batch normalizations between layers. The models were trained for the final transpose layer to reconstruct the original signal with the loss function logarithmic scaled absolute voltage weighted root mean square error and KL loss (β) set to 0. The models were trained using Adam optimization, a learning rate of 0.00001, and were trained for approx. 100 epochs with a batch size of 32 on 1.18 million ECGs.

### Data splitting

Each participant had a single sleep study included for analysis. The sleep study was paired with one or more ECGs available within 1 year before or after. A random selection of 10% were held out for testing and reporting, and the remainder 90% were utilized for training (**Figure 1**). The training dataset was partitioned using a stratified 5-fold cross validation scheme and a reduced training dataset limited to a single temporally most proximate ECG per sleep study was used. Feature selection and hyperparameter tuning were performed separately in each of the 5 folds and selected based on best cross-validation performance (i.e., area under the receiver operating characteristics curve [AUROC]). Then the final models (clinical, ECG, and combined) were trained on the full training dataset. The final models’ performance was then tested and reported from the 10% held-out test set.

**Figure 1.**
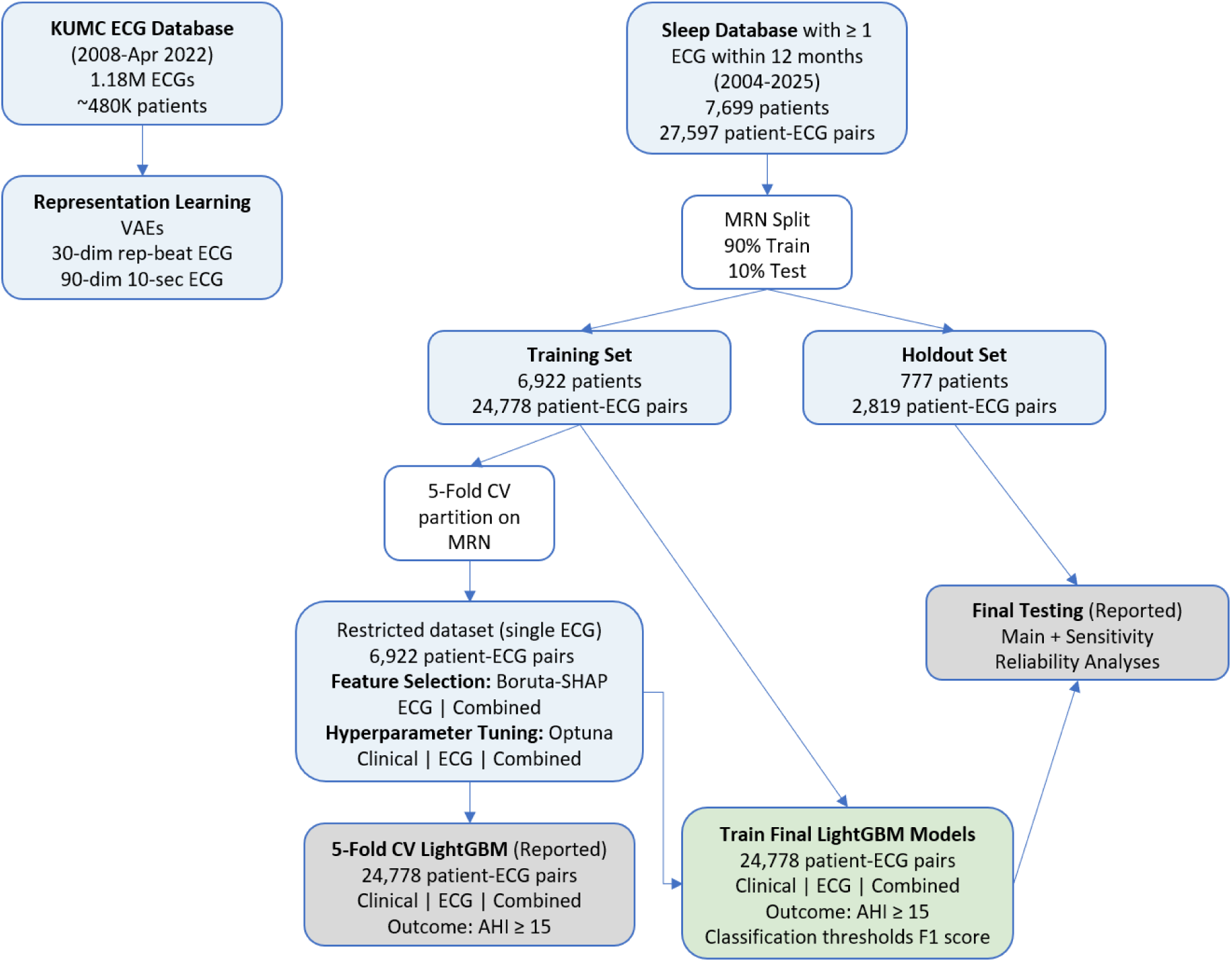
Study overview and sample selection flowchart.

**Figure 2.**
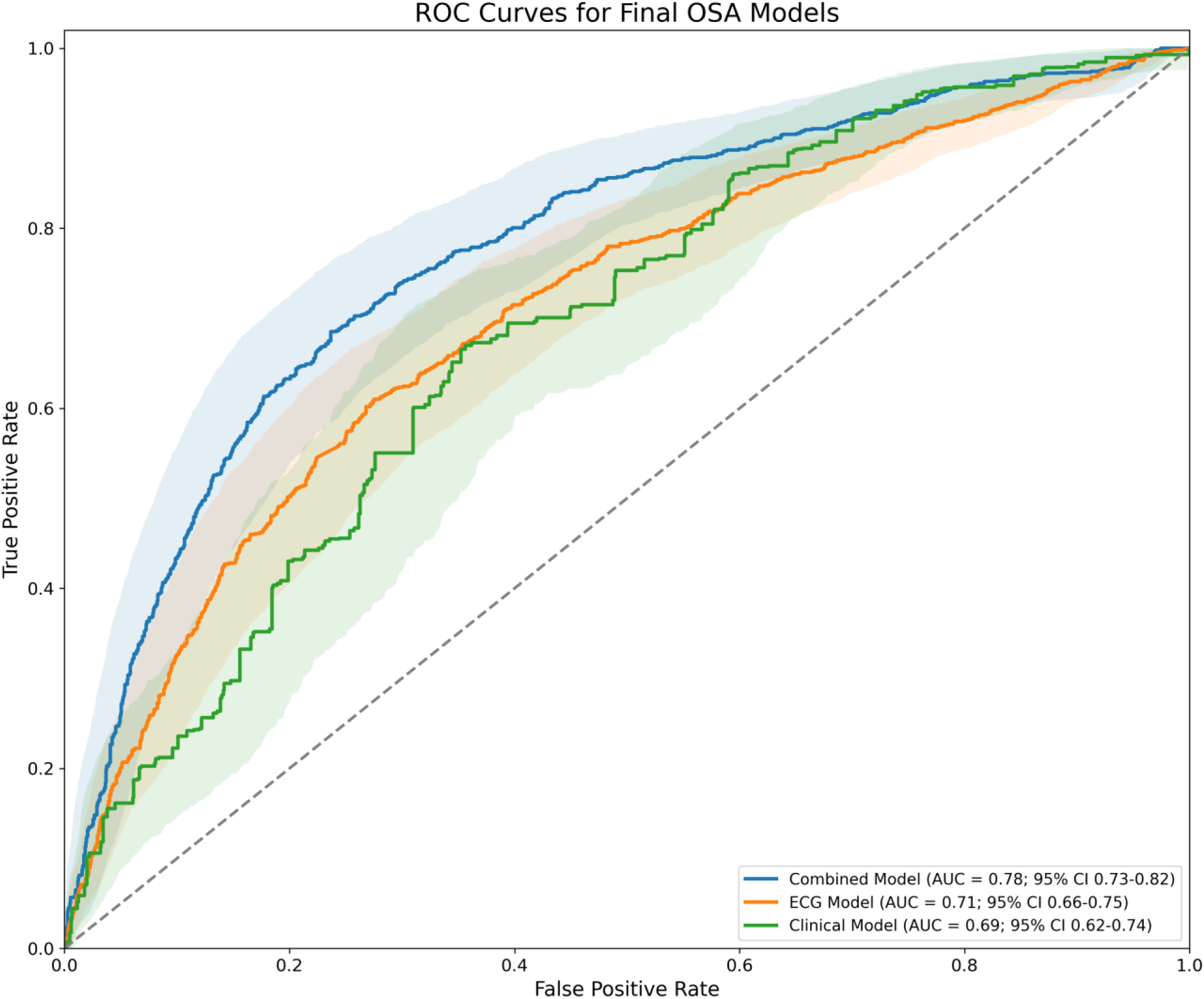
Receiver-operating characteristic (ROC) curves in the primary holdout test set analysis (n = 777 patients, 2,899 ECGs).

### Machine learning models

Three LightGBM models were trained to predict moderate-to-severe OSA: (1) six clinical features only (age, sex, weight, height, BMI, ESS); (2) ECG features only (all summary ECG metrics and VAE latent dimensions); and (3) a combined model incorporating all features. LightGBM is a highly efficient histogram-based gradient boosting framework well suited for complex tabular clinical data [25]. Models were optimized using AUROC as the training metric, with a learning rate of 0.01 and early stopping if cross-validation performance did not improve over 1,000 boosting rounds. Class weights were balanced considering the ∼1:2 imbalance between moderate-to-severe and no-to-mild OSA. Feature selection and hyperparameter tuning were performed on the reduced training subset limited to one sleep study-ECG pair per patient, chosen by closest temporal proximity. Within each of 5 stratified cross-validation folds (80% training and 20% validation, partitioned by patient), features were selected (except clinical model where all 6 features were retained) using Boruta with SHAP importance (80 trials) [26] and hyperparameters were tuned using Bayesian optimization (Optuna, 200 trials) [27], separately for each of the three models. The feature set and hyperparameters from the fold with the best validation AUROC were carried forward. Final models were trained on the full training dataset using the selected features and hyperparameters, with prediction thresholds chosen to maximize the F1-score. The three final models were evaluated on the independent 10% holdout test set that was not used for any training, tuning, or feature-selection step, with AUROC and 95% confidence intervals obtained via bootstrapping (1,000 iterations). Models were evaluated in the full holdout set (main analysis, ≥1 ECG per sleep study) and in two additional sensitivity analyses: (1) restricted to a single, most temporally proximate ECG per sleep study, and (2) restricted to ECGs acquired within 30 days of the sleep study. Among the holdout patients with more than one ECG, inter-ECG reliability of model predictions was assessed using Fleiss’ kappa.

### Feature importance

Feature importance was assessed using two complementary LightGBM metrics: information gain (average improvement in model prediction at each split) and split count (proportion of decision tree splits using each feature). Both metrics were obtained from the model trained on the entire training dataset. Because feature selection differed across the clinical, ECG, and combined models, feature importance is reported separately for each model. Latent conditional traversal was also performed to visualize the ECG morphological variation encoded by the important VAE features. We systematically varied the VAE latent variable across ±2 standard deviations in the KUMC ECG database (>1 million ECGs) while holding all others latent variables at their median, then decoding the ECG waveforms to characterize the morphological variation encoded.

### Statistical analysis

Continuous variables are presented as mean ± standard deviation; categorical variables as N (%). Differences between OSA severity groups were assessed using Student’s t-test for continuous variables and χ² test for categorical variables. Model performance was compared by AUROC. Accuracy, F1-score, precision, sensitivity (recall), and specificity, computed at the F1-score-optimized prediction threshold, were reported as secondary metrics for the main holdout analysis. A p-value threshold of <0.05 was considered statistically significant. AUROC 95% confidence intervals were obtained via bootstrapping (1,000 iterations). Inter-ECG reliability among holdout patients with repeated ECGs was assessed using Fleiss’ kappa and percent observed agreement; model performance using the majority-vote prediction across repeated ECGs was also reported.

## Results

### Sample characteristics

The dataset included 7,699 patients with an index sleep study and 27,597 ECGs (**Figure 1**). A total of 534 patients (6.9%) also had more than one sleep study. The sample was 52.2% women (N=4,013), with mean ± SD age 58.0±14.2 years, BMI 33.6±8.2 kg/m^²^, and ESS score 8.8±5.1. Among all participants, 36.5% (N=2,806) met criteria for moderate-to-severe OSA (AHI ≥15 events/h) and 63.5% (N=4,893) were classified as no or mild OSA. The mean ± SD absolute time between the diagnostic sleep study and ECG acquisition was 143.7±107.7 days. Participant characteristics are presented in **Table 1**.

**Table 1.** Participant characteristics by OSA severity group.

| **Variable** | **Total**<br>**(N = 7,699)** | **No or mild**<br>**OSA, AHI<15**<br>**(N=4,893)** | **Moderate or severe**<br>**OSA, AHI≥15**<br>**(N=2,806)** | **p** |
| --- | --- | --- | --- | --- |
| Age, years | 58.0 ± 14.2 | 56.6 ± 14.6 | 60.5 ± 13.0 | <0.001 |
| Sex, female, N (%) | 4013 (52.2%) | 2783 (56.9%) | 1230 (43.9%) | <0.001 |
| BMI, kg/m<sup>2</sup> | 33.6 ± 8.2 | 32.6 ± 7.8 | 35.4 ± 8.7 | <0.001 |
| Weight, kg | 98.0 ± 25.4 | 94.2 ± 23.5 | 104.6 ± 27.1 | <0.001 |
| Height, cm | 170.8 ± 10.8 | 170.2 ± 10.9 | 171.9 ± 10.6 | <0.001 |
| ESS score | 8.8 ± 5.1 | 8.8 ± 5.1 | 8.8 ± 5.0 | 0.769 |
| Race: White, N (%) | 4538 (58.9%) | 3293 (67.3%) | 1245 (44.4%) | <0.001 |
| Absolute days between<br>ECG and sleep study | 143.7 ± 107.7 | 143.6 ± 107.7 | 144.0 ± 107.7 | 0.859 |
| In-lab PSG, N (%) | 2894 (37.6%) | 2394 (48.9%) | 500 (17.8%) | <0.001 |
| AHI, events/h | 16.4 ± 18.6 | 5.7 ± 4.2 | 35.0 ± 19.2 | <0.001 |

Abbreviations: AHI, apnea-hypopnea index; BMI, body mass index; ESS, Epworth Sleepiness Scale score; OSA: obstructive sleep apnea; PSG, polysomnography; SD: standard deviation. Values are mean ± SD or N (%).

### Combined model had best performance predicting moderate-to-severe OSA

The combined model achieved the best cross-validation performance among the three assessed models (**Table 2**). Average 5-fold cross-validation AUROC (95% CI) was 0.713 (0.661, 0.760) for the combined model, compared to 0.648 (0.590, 0.702; p<0.001) for the clinical-only model and 0.663 (0.609, 0.721; p<0.001) for the ECG-only model. In the independent holdout test set (main analysis, ≥1 ECG per sleep study), holdout AUROC was 0.776 (0.727, 0.822) for the combined model, as compared to 0.686 (0.632, 0.737; p<0.001) for the clinical-only model, and 0.708 (0.661, 0.752; p<0.001) for the ECG-only model, with accuracy (at the F1-optimized classification threshold) of 0.711, 0.619, and 0.655, respectively. Restricting to a single, most temporally proximate ECG per sleep study (N=777) and to ECGs acquired within 30 days of the sleep study (437 ECGs from 270 patients) yielded similar holdout AUROCs (**Table 2**). Among the 488 of 777 holdout patients with more than one ECG, measures for inter-ECG reliability of model predictions yielded 89% agreement with Fleiss’s κ=0.73 while the combined model had 96% agreement with κ=0.90.

**Table 2.** Areas under receiver-operating characteristic curves (AUROCs) with 95% confidence intervals for Light Gradient-Boosted Machine models.

| Data split | N<br>(patients / ECGs) | Clinical model | ECG model | Combined model |
| --- | --- | --- | --- | --- |
| **Cross-validation (5-fold)** | **6,922 / 24,778** |  |  |  |
| Best fold | - | 0.670<br>(0.624, 0.713) | 0.693<br>(0.651, 0.735) | 0.731<br>(0.688, 0.767) |
| Average of 5 folds | - | 0.648<br>(0.590, 0.702) | 0.663<br>(0.609, 0.721) | 0.713<br>(0.661, 0.760) |
| **Holdout set** | **777 / 2,819** |  |  |  |
| Main analysis | 777 / 2,819 | 0.686<br>(0.632, 0.737) | 0.708<br>(0.661, 0.752) | 0.776<br>(0.727, 0.822) |
| Single ECG per sleep study | 777 / 777 | 0.691<br>(0.654, 0.728) | 0.693<br>(0.649, 0.729) | 0.758<br>(0.721, 0.792) |
| ECGs within 30 days of sleep study | 270 / 437 | 0.659<br>(0.573, 0.736) | 0.672<br>(0.586, 0.746) | 0.767<br>(0.691, 0.836) |

Abbreviations. ECG: electrocardiogram.

**Table 3.**
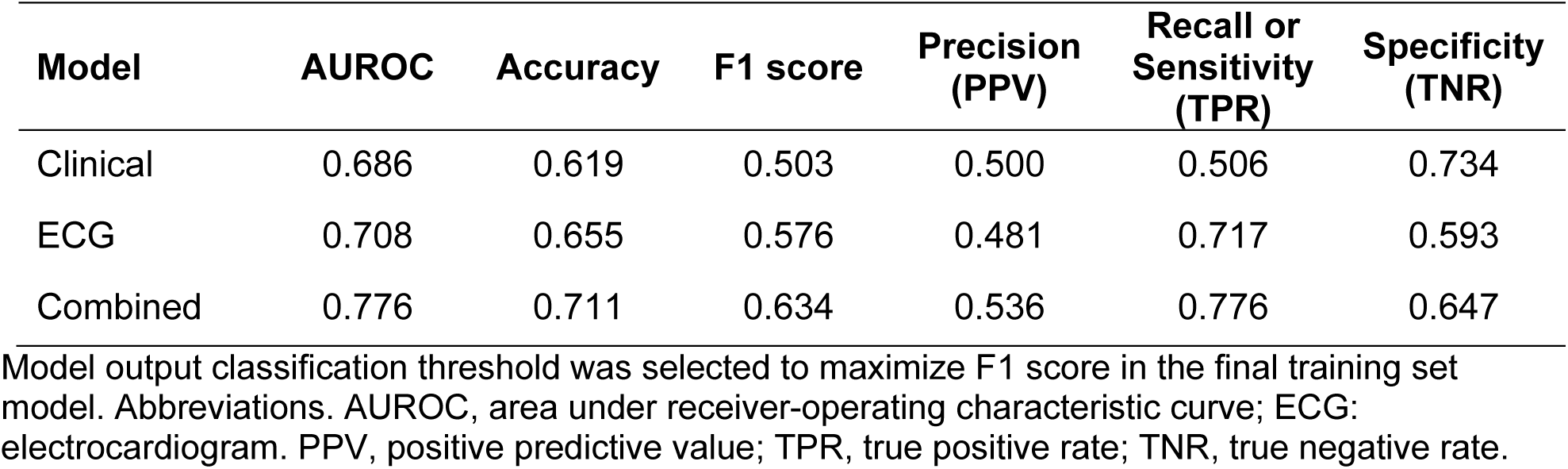
Performance metrics of the three Light Gradient-Boosted Machine models in the primary holdout test set analysis (n = 777 patients, 2,809 ECGs).

| Model | AUROC | Accuracy | F1 score | Precision (PPV) | Recall or Sensitivity (TPR) | Specificity (TNR) |
| --- | --- | --- | --- | --- | --- | --- |
| Clinical | 0.686 | 0.619 | 0.503 | 0.500 | 0.506 | 0.734 |
| ECG | 0.708 | 0.655 | 0.576 | 0.481 | 0.717 | 0.593 |
| Combined | 0.776 | 0.711 | 0.634 | 0.536 | 0.776 | 0.647 |

Model output classification threshold was selected to maximize F1 score in the final training set model. Abbreviations. AUROC, area under receiver-operating characteristic curve; ECG: electrocardiogram. PPV, positive predictive value; TPR, true positive rate; TNR, true negative rate.

### Feature importance analysis

**Figure 3** shows feature importance (gain and split) separately for the clinical, ECG, and combined models, reflecting the features retained by Boruta-SHAP selection for each model. In the clinical model, BMI, weight, and age were the top predictors, consistent with the role of adiposity and demographic risk in OSA. In the ECG-only model, representative-beat VAE embeddings (latent variables 5, 1, and 25) were the top predictors, alongside conventional ECG measures including aVF and V1 R-wave amplitude, V2 QRS intrinsicoid deflection, and PR interval. In the combined model, age, BMI, and weight remained the top predictors along with representative-beat VAE latent variables 29, 1, 5, and 25, height and sex. Additionally, the p-wave voltage-time integral in the reconstructed orthogonal X lead and four latent variables from the 10-second VAE were also among the top predictors.

**Figure 3.**
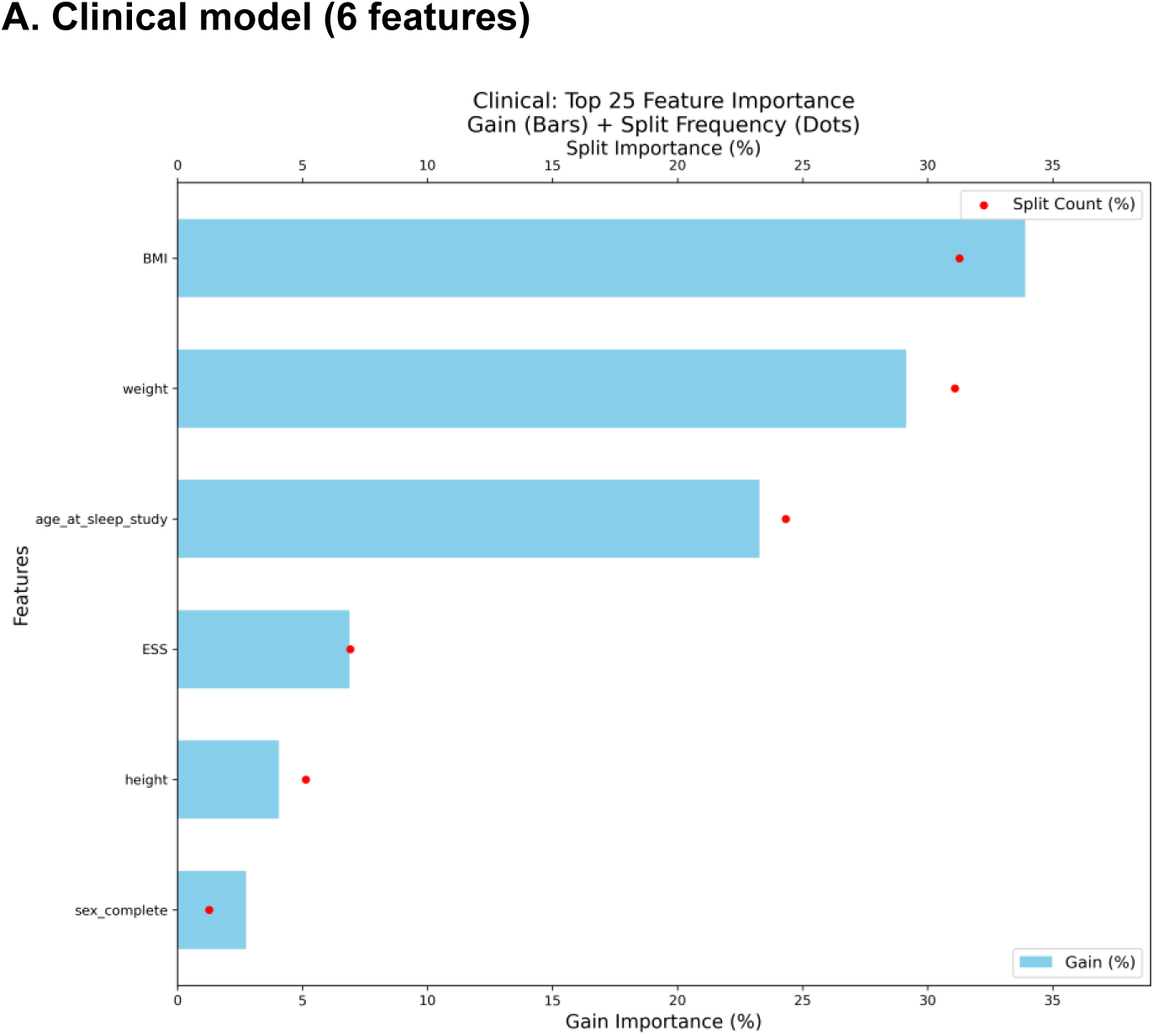
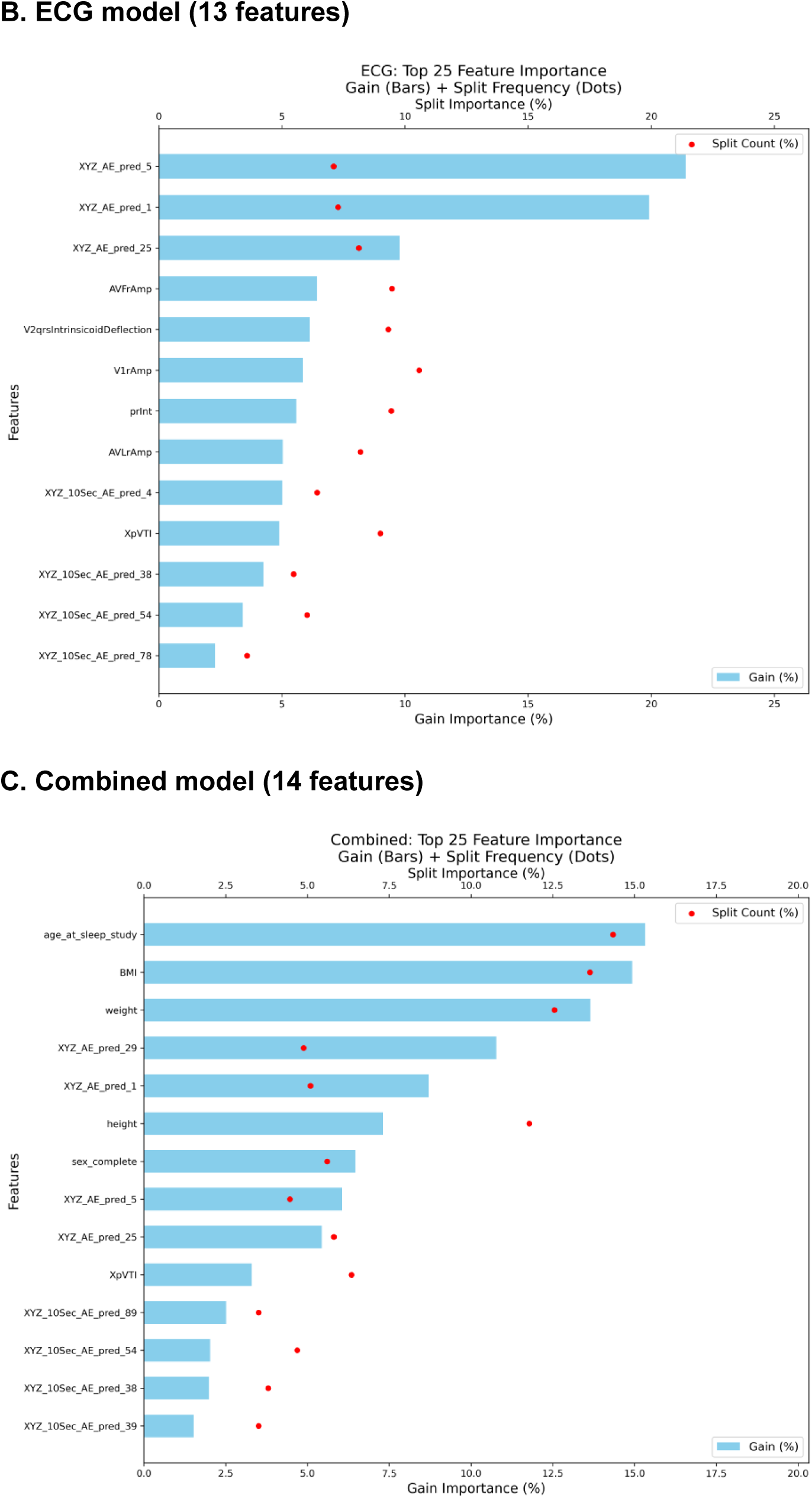
Feature importance (gain and split) for the (A) clinical, (B) ECG, and (C) combined models, based on the features selected by Boruta-SHAP for each model.

### Latent ECG representation analysis

Latent conditional traversal to visualize the morphological variation encoded by the highest-importance representative-beat VAE embeddings (1, 5, 25, and 29) is shown in **Figure 4**. Systematically varying these latent variables seemed to modulate specific ECG morphological characteristics.

**Figure 4.**
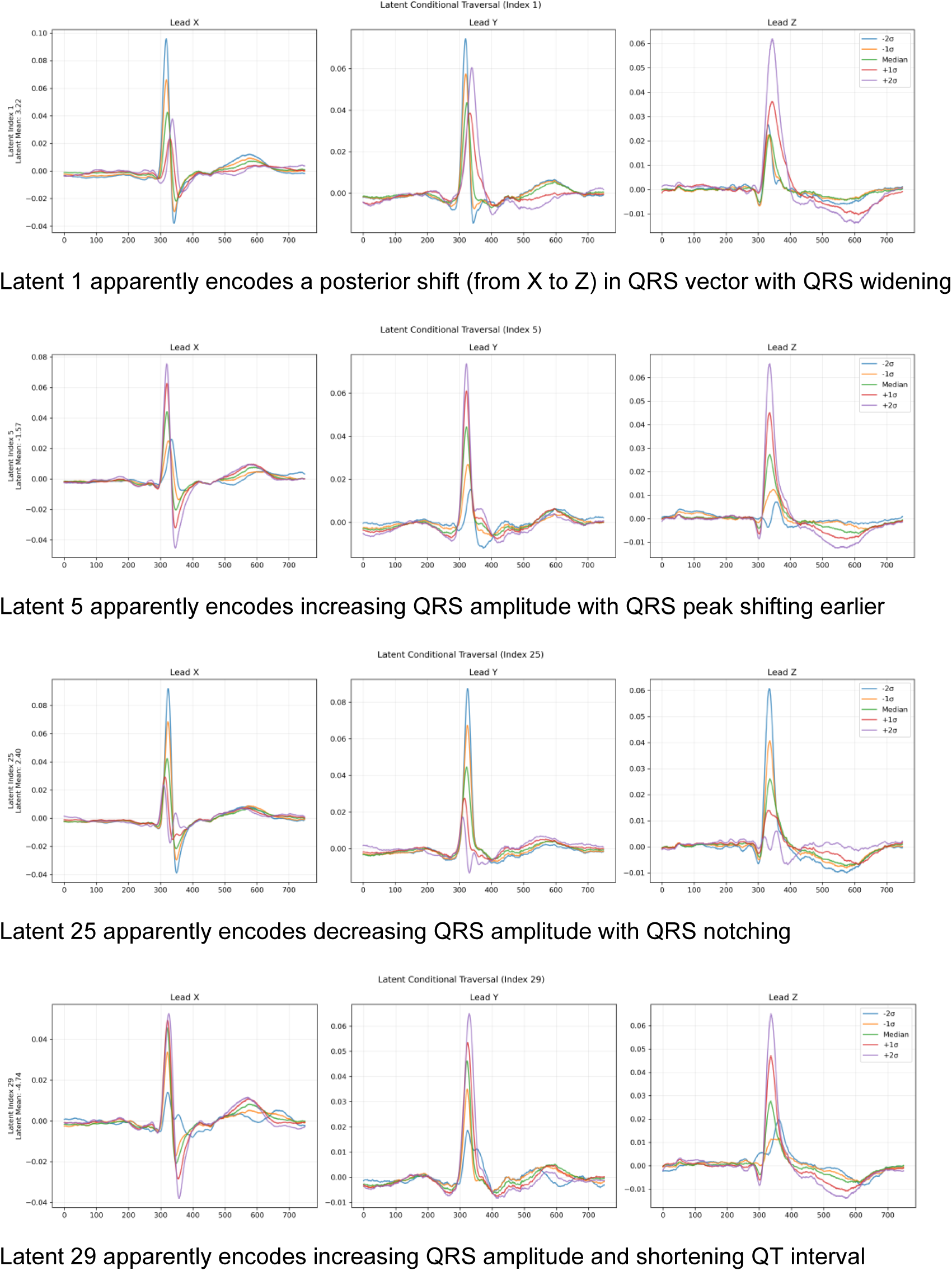
Latent conditional traversal for representative-beat VAE latent variables 1, 5, 25, and 29 with reconstructed X-, Y-, and Z-leads. ECGs are reconstructed at ±2 SD, ±1SD, and median, with all other latent variables fixed at their median values.

## Discussion

In this large, single-center, retrospective study, we demonstrated that ECG features derived from standard 12-lead recordings using a pretrained VAE improve prediction of moderate-to-severe OSA when combined with established clinical factors. The combined model achieved 5-fold cross-validation average AUROC of 0.71 (95% C.I. 0.66, 0.76) and holdout test AUROC of 0.78 (95% C.I. 0.73, 0.82), outperforming the clinical-only and ECG-only models and comparing favorably with prior machine learning based OSA prediction studies [28–31]. Feature importance analysis showed that BMI and weight drove the clinical model, consistent with the established role of adiposity in OSA pathophysiology, while VAE-derived latent ECG embeddings outweighed conventional ECG measurements, with the most informative conventional features suggesting left ventricular hypertrophy (LVH), atrial remodeling, and autonomic dysregulation as being associated with OSA.

The potential of using predictive models to identify OSA, either from demographics and symptom data, or additionally incorporating clinical data has been extensively explored. Our prior study in a 17,448-participant multi-center cohort applied machine learning using age, sex, BMI, and race to predict OSA and reported validation AUROCs of 0.68 [32]. In the current study, the clinical-only model yielded a comparable cross-validation AUROC at 0.65 (holdout set 0.69), and the addition of ECG features improved cross-validation AUROC to 0.71 (holdout set 0.78), a clinically meaningful increment. Importantly, performance gains were consistent in all sensitivity analyses, including restricting to a single ECG per participant, and to ECGs that were only obtained within 30 days of the index sleep study. These findings align with the broader machine learning literature on OSA screening. For instance, a support vector machine model trained on demographic and clinical features alone in a large clinic-based cohort achieved an AUROC of 0.82 for detecting OSA [33], comparable in magnitude to the clinical-feature gains we observed when adding ECG-derived features. More broadly, a systematic review of 63 studies applying machine learning to OSA diagnosis reported best-case AUROCs as high as 0.97 for internally validated models and 0.98 for externally validated models [34]. However, this review also highlights that these somewhat optimistic estimates were heavily dependent on model type and feature set, though most studies lacked external validation in diverse cohorts. Similarly, our study does not report on an external validation set. Conversely, we included a 10% holdout set to mitigate this limitation, but we recognize that external validation of our models is warranted.

Importantly, the feature importance analysis in our reported clinical model revealed biologically interpretable patterns. BMI and weight accounted for most of the gain importance, reflecting the central mechanistic role of excess adiposity in OSA through pharyngeal fat deposition, reduced lung volumes [35–37], and impaired upper airway muscle function [38,39]. However, the modest performance of the clinical model underscores the well-recognized limitation that OSA is phenotypically heterogeneous [40], and many individuals with moderate-to-severe disease may not fit the expected anthropometric profile determined by factors such as age, sex, and BMI. Our seminal study assessing the clinical surrogates of symptom heterogeneity demonstrated that OSA symptom subtypes predict cardiovascular outcomes differentially [41], illustrating that risk factor-based tools alone are insufficient to capture the full clinical spectrum of OSA. Notably, sleepiness, based on the ESS score, showed a minimal relative importance in our clinical model, consistent with the known dissociation between subjective sleepiness and objective OSA severity in clinical populations [42,43]. Despite this, ESS remained among the top clinical predictors in the machine learning models, possibly because it is likely associated with the other predictors in non-linear ways that could be captured by tree-based models.

More recently, other data modalities are increasingly being assessed to improve OSA prediction. In the ECG domain, most deep learning work has applied single-lead ECG signals recorded during polysomnography to detect discrete apnea or hypopnea events at high temporal resolution, a task distinct from diagnosing OSA at the whole-night level [18]. Other studies have instead used sleep study ECG signals for OSA screening itself, achieving high sensitivity and specificity relative to the AHI-based gold standard [44]. Fewer studies still have applied deep learning to a standalone clinical 12-lead ECG obtained independently of a sleep study, as in a model trained on 11,299 patients that achieved an AUC of 0.80 for OSA diagnosis [14]. In our current study, the ECG-only model achieved OSA classification performance on par with or exceeding the clinical model. It is also noteworthy that representative-beat VAE embeddings (5, 1, and 25) had higher feature importance than conventional ECG measurements. Amplitude of R wave in leads aVF, V1, and aVL were selected suggesting correlation of LVH with OSA. This is biologically plausible, as OSA promotes ventricular remodeling through chronic sympathetic activation, intermittent hypoxemia, and repetitive increases in right ventricular afterload [45,46], which may lead to systemic hypertension [47], a condition that is highly prevalent in OSA and that also independently promotes LVH [48]. The selection of P-wave and PR-interval features may reflect atrial remodeling and autonomic dysregulation associated with OSA [49,50].

Our primary findings are that the combined model outperformed both the clinical and ECG models. In addition to clinical features of age, BMI, weight, height, and sex, the other important features were representative-beat as well as full 10-second ECG VAEs. The only non-VAE feature included was P-wave voltage-time integral in X-lead reflecting a potential role for atrial remodeling. The most important VAE features in the combined model were the representative beat latent variables 29, 1, 5, and 25. The latent traversal analysis confirmed that the top VAE features may encode meaningful morphological variation. Latent 1 appeared to affect posterior shift in QRS along with QRS widening or development of left bundle branch block. Latent variables 5 and 25 affected QRS amplitude and morphology. Low values of latent 29 appeared to reduce QRS voltage and resulted in QT interval prolongation and T wave abnormality. These suggest the VAE embeddings capture phenotypes such as ventricular geometry and repolarization that are consistent with autonomic dysregulation and arrhythmia risk consistent with prior literature [51], as well as effects of adiposity on ECG [52], with plausible mechanistic links to OSA-related cardiac remodeling.

It is important to recognize that patients predicted by our ECG-based models to have OSA, particularly those with high representative-beat VAE feature importance suggestive of LVH and atrial remodeling, may already experience established structural cardiac abnormalities detectable by echocardiography. This is consistent with a meta-analysis of echocardiographic studies showing a pooled LVH prevalence of 45% in OSA and an increased risk of LVH with greater OSA severity [53]. A broader systematic review and meta-analysis of imaging studies similarly found that, compared with controls, patients with OSA had significantly wider left atrial diameter, greater left ventricular mass, and reduced left ventricular ejection fraction [54]. These structural changes appear to be reversible, as LVH has been shown to regress with continuous positive airway pressure therapy [55]. Given the overlap between the cardiac phenotypes implicated by our ECG-derived features and those documented using echocardiography in OSA, future work linking available echocardiographic data in our cohort could help quantify and validate the cardiac-structural correlates underlying our reported predictions.

This study also contributes to a growing literature on ECG- and wearable-based OSA screening. Apple Watch recently received FDA clearance for a sleep apnea notification algorithm [56], and photoplethysmography-based wearables are increasingly evaluated for automated OSA detection [57]. Our results complement these developments by demonstrating that a single standard 12-lead ECG already acquired routinely in primary care contains independent predictive information about OSA. This is particularly relevant given the high prevalence of OSA among patients in cardiovascular clinics who already obtain 12-lead ECGs and the opportunity to screen for OSA at the point of ECG acquisition without additional patient burden.

Our study includes several meaningful strengths such as the large sample size (N = 7,699), standardized ECG processing, use of diverse feature types (conventional morphological features, and two independent VAE-derived representations), and a robust machine learning pipeline with explicit holdout validation and sensitivity analyses. However, important limitations include a single-center retrospective design requiring external validation, limited racial/ethnic diversity of the study population, variability between ECG and sleep study assessment timing, and absence of additional OSA risk factors (neck circumference, snoring history, craniofacial anatomy). Moreover, the absolute performance of the combined model (cross-validation AUROC 0.71; holdout AUROC 0.78), which, while promising, suggests that additional sources of heterogeneity such as comorbidity burden or nocturnal oxygen desaturation profiles, may be needed to achieve clinical-grade prediction performance. It is notable that the holdout analysis outperformed our average cross-validation AUROC. We took extra care to ensure there was no data leakage, and the holdout test set was not used for any other steps except for testing and reporting the performance of the final model derived in the training set. This higher AUROC may be due to random chance, though the final model trained using all of the training data could conceivably be better than the cross-fold models trained only 80% of the training data.

In conclusion, ECG features derived from standard 12-lead recordings improved prediction of moderate-to-severe OSA when combined with clinical factors, achieving cross-validation AUROC of 0.71 (95% CI 0.66-0.76), with holdout AUROC (0.78, 95% CI 0.73-0.82) and comparable performance when ECG acquisition was temporally proximate to the sleep study. Feature importance analyses identified biologically plausible clinical and ECG predictors, including markers of left ventricular hypertrophy, atrial conduction (P-wave/PR-interval) features, and latent VAE-derived representations of ventricular repolarization, consistent with cardiovascular sequelae relevant to OSA. ECG may be useful as a screening adjunct for OSA in cardiology and potentially in primary care settings. Prospective validation in diverse populations and investigation of whether ECG-based screening predicts cardiovascular outcomes in OSA are still needed.

## Data Availability

All data produced in the present study are not available for sharing, as it represents data from medical records in an academic medical institutions. However, the authors encourage collaboration requests to use the data.

## References

1. Benjafield AV, Ayas NT, Eastwood PR, et al. Estimation of the global prevalence and burden of obstructive sleep apnoea: a literature-based analysis. Lancet Respir Med. Aug 2019;7(8):687–698. doi:10.1016/S2213-2600(19)30198-5

2. Lévy P, Kohler M, McNicholas WT, et al. Obstructive sleep apnoea syndrome. Nature Reviews Disease Primers. 2015;1(1)doi:10.1038/nrdp.2015.15

3. Yeghiazarians Y, Jneid H, Tietjens JR, et al. Obstructive Sleep Apnea and Cardiovascular Disease: A Scientific Statement From the American Heart Association. Circulation. 2021;144(3)doi:10.1161/cir.0000000000000988

4. Kohler M, Stradling JR. Mechanisms of vascular damage in obstructive sleep apnea. Nature Reviews Cardiology. 2010;7(12):677–685. doi:10.1038/nrcardio.2010.145

5. McEvoy RD, Antic NA, Heeley E, et al. CPAP for Prevention of Cardiovascular Events in Obstructive Sleep Apnea. New England Journal of Medicine. 2016;375(10):919–931. doi:10.1056/NEJMoa1606599

6. Barbé F, Durán-Cantolla J, Sánchez-de-la-Torre M, et al. Effect of Continuous Positive Airway Pressure on the Incidence of Hypertension and Cardiovascular Events in Nonsleepy Patients With Obstructive Sleep Apnea. Jama. 2012;307(20)doi:10.1001/jama.2012.4366

7. Pack AI, Magalang UJ, Singh B, Kuna ST, Keenan BT, Maislin G. Randomized clinical trials of cardiovascular disease in obstructive sleep apnea: understanding and overcoming bias. Sleep. 2021;44(2)doi:10.1093/sleep/zsaa229

8. Xie C, Zhu R, Tian Y, Wang K. Association of obstructive sleep apnoea with the risk of vascular outcomes and all-cause mortality: a meta-analysis. BMJ Open. 2017;7(12)doi:10.1136/bmjopen-2016-013983

9. Zhang X, Fan J, Guo Y, et al. Association between obstructive sleep apnoea syndrome and the risk of cardiovascular diseases: an updated systematic review and dose–response meta-analysis. Sleep Medicine. 2020;71:39–46. doi:10.1016/j.sleep.2020.03.011

10. Nagappa M, Liao P, Wong J, et al. Validation of the STOP-Bang Questionnaire as a Screening Tool for Obstructive Sleep Apnea among Different Populations: A Systematic Review and Meta-Analysis. PLoS One. 2015;10(12):e0143697. doi:10.1371/journal.pone.0143697

11. Netzer NC, Stoohs RA, Netzer CM, Clark K, Strohl KP. Using the Berlin Questionnaire to identify patients at risk for the sleep apnea syndrome. Ann Intern Med. Oct 5 1999;131(7):485–91. doi:10.7326/0003-4819-131-7-199910050-00002

12. Chiu HY, Chen PY, Chuang LP, et al. Diagnostic accuracy of the Berlin questionnaire, STOP-BANG, STOP, and Epworth sleepiness scale in detecting obstructive sleep apnea: A bivariate meta-analysis. Sleep Med Rev. Dec 2017;36:57–70. doi:10.1016/j.smrv.2016.10.004

13. Fraley MA, Birchem JA, Senkottaiyan N, Alpert MA. Obesity and the electrocardiogram. Obes Rev. Nov 2005;6(4):275–81. doi:10.1111/j.1467-789X.2005.00199.x

14. Covassin N, Liu K, Bukartyk J, et al. Deep Neural Network Algorithm Using the Electrocardiogram for Detection of Obstructive Sleep Apnea. JACC: Advances. 2025;4(10)doi:10.1016/j.jacadv.2025.102139

15. Roche F, Roux M, Benali K, Pichot V. Electrocardiographic assessment in obstructive sleep apnea: bridging pathophysiology and clinical practice. Frontiers in Physiology. 2026;17doi:10.3389/fphys.2026.1777406

16. Qin H, Keenan BT, Mazzotti DR, et al. Heart rate variability during wakefulness as a marker of obstructive sleep apnea severity. Sleep. 2021;44(5)doi:10.1093/sleep/zsab018

17. Wang Z, Jiang F, Xiao J, et al. Heart rate variability changes in patients with obstructive sleep apnea: A systematic review and meta-analysis. Journal of Sleep Research. 2022;32(1)doi:10.1111/jsr.13708

18. Zarei A, Beheshti H, Asl BM. Detection of sleep apnea using deep neural networks and single-lead ECG signals. Biomedical Signal Processing and Control. 2022;71doi:10.1016/j.bspc.2021.103125

19. Erdenebayar U, Kim YJ, Park J-U, Joo EY, Lee K-J. Deep learning approaches for automatic detection of sleep apnea events from an electrocardiogram. Computer Methods and Programs in Biomedicine. 2019;180doi:10.1016/j.cmpb.2019.105001

20. Gupta A, Harvey CJ, DeBauge A, et al. Machine learning to classify left ventricular hypertrophy using electrocardiographic feature extraction by variational autoencoder. Heart Rhythm O2. Jul 2026;7(7):1324–1334. doi:10.1016/j.hroo.2026.03.019

21. Harvey C, Shomaji S, Yao Z, Noheria A. ECG Latent Feature Extraction with Autoencoders for Downstream Prediction Tasks. presented at: 2025 IEEE Signal Processing in Medicine and Biology Symposium (SPMB); 2025;

22. Berry RB, Brooks R, Gamaldo C, et al. AASM Scoring Manual Updates for 2017 (Version 2.4). J Clin Sleep Med. May 15 2017;13(5):665–666. doi:10.5664/jcsm.6576

23. Johns MW. A new method for measuring daytime sleepiness: the Epworth sleepiness scale. Sleep. Dec 1991 1991;14:540–545.

24. Kors JA, van Herpen G, Sittig AC, van Bemmel JH. Reconstruction of the Frank vectorcardiogram from standard electrocardiographic leads: diagnostic comparison of different methods. Eur Heart J. Dec 1990;11(12):1083–92. doi:10.1093/oxfordjournals.eurheartj.a059647

25. Yıldız AY, Kalayci A. Gradient Boosting Decision Trees on Medical Diagnosis Over Tabular Data. presented at: 2025 IEEE International Conference on AI and Data Analytics (ICAD); 2025;

26. BorutaShap: A wrapper feature selection method which combines the Boruta feature selection algorithm with Shapley values. . Version Version 1.1. 2020.

27. Akiba T, Sano S, Yanase T, Ohta T, Koyama M. Optuna. presented at: Proceedings of the 25th ACM SIGKDD International Conference on Knowledge Discovery & Data Mining; 2019;

28. Hwang N, Westover MB, Mazzotti D, et al. 0509 Applying Machine Learning to Predict Obstructive Sleep Apnea Using Electronic Health Records. Sleepj. 2026;49(Supplement_1):A227–A227. doi:10.1093/sleep/zsag091.0508

29. Liu K, Geng S, Shen P, Zhao L, Zhou P, Liu W. Development and application of a machine learning-based predictive model for obstructive sleep apnea screening. Frontiers in Big Data. 2024;7doi:10.3389/fdata.2024.1353469

30. Dai R, Yang K, Zhuang J, et al. Enhanced machine learning approaches for OSA patient screening: model development and validation study. Scientific Reports. 2024;14(1)doi:10.1038/s41598-024-70647-5

31. Yan X, Wang L, Liang C, et al. Development and assessment of a risk prediction model for moderate-to-severe obstructive sleep apnea. Frontiers in Neuroscience. 2022;16doi:10.3389/fnins.2022.936946

32. Holfinger SJ, Lyons MM, Keenan BT, et al. Diagnostic Performance of Machine Learning-Derived OSA Prediction Tools in Large Clinical and Community-Based Samples. Chest. Mar 2022;161(3):807–817. doi:10.1016/j.chest.2021.10.023

33. Lai F, Chiang AA, Liu Y-T, Lee P-L, Huang W-C. Support vector machine prediction of obstructive sleep apnea in a large-scale Chinese clinical sample. Sleep. 2020;43(7)doi:10.1093/sleep/zsz295

34. Ferreira-Santos D, Amorim P, Silva Martins T, Monteiro-Soares M, Pereira Rodrigues P. Enabling Early Obstructive Sleep Apnea Diagnosis With Machine Learning: Systematic Review. Journal of Medical Internet Research. 2022;24(9)doi:10.2196/39452

35. Baker E, Chanamolu M, Nieri C, White SF, Brandt J, Gillespie MB. The Effect of Tongue Volume and Adipose Content on Obstructive Sleep Apnea: Meta-analysis & Systematic Review. OTO Open. 2025;9(2)doi:10.1002/oto2.70067

36. Isono S. Obesity and obstructive sleep apnoea: Mechanisms for increased collapsibility of the passive pharyngeal airway. Respirology. 2011;17(1):32–42. doi:10.1111/j.1440-1843.2011.02093.x

37. Schwartz AR, Patil SP, Squier S, Schneider H, Kirkness JP, Smith PL. Obesity and upper airway control during sleep. Journal of Applied Physiology. 2010;108(2):430–435. doi:10.1152/japplphysiol.00919.2009

38. D’Angelo GF, de Mello AAF, Schorr F, et al. Muscle and visceral fat infiltration: A potential mechanism to explain the worsening of obstructive sleep apnea with age. Sleep Medicine. 2023;104:42–48. doi:10.1016/j.sleep.2023.02.011

39. Shi L, Wang H, Wei L, Hong Z, Wang M, Wang Z. Pharyngeal constrictor muscle fatty change may contribute to obstructive sleep apnea-hypopnea syndrome: a prospective observational study. Acta Oto-Laryngologica. 2016;136(12):1285–1290. doi:10.1080/00016489.2016.1205220

40. Zinchuk A, Yaggi HK. Phenotypic Subtypes of OSA. Chest. 2020;157(2):403–420. doi:10.1016/j.chest.2019.09.002

41. Mazzotti DR, Keenan BT, Lim DC, Gottlieb DJ, Kim J, Pack AI. Symptom Subtypes of Obstructive Sleep Apnea Predict Incidence of Cardiovascular Outcomes. Am J Respir Crit Care Med. Aug 15 2019;200(4):493–506. doi:10.1164/rccm.201808-1509OC

42. Frangopoulos F, Zannetos S, Nicolaou I, et al. The Complex Interaction Between the Major Sleep Symptoms, the Severity of Obstructive Sleep Apnea, and Sleep Quality. Frontiers in Psychiatry. 2021;12doi:10.3389/fpsyt.2021.630162

43. Sartori G, Di Chiara C, Gretter A, Fantin A, Crisafulli E. Cancer History and Subjective Sleepiness in Obstructive Sleep Apnea: A Real-World Observational Study. Journal of Clinical Medicine. 2026;15(12)doi:10.3390/jcm15124573

44. Nygate Y, Sprague M, Rusk S, Fernandez C, Watson N. 0680 Clinical Validation of ECG-Based Obstructive Sleep Apnea Screening Using Machine Learning. Sleep. 2025;48(Supplement_1):A296–A296. doi:10.1093/sleep/zsaf090.0680

45. DiCaro MV, Lei K, Yee B, Tak T. The Effects of Obstructive Sleep Apnea on the Cardiovascular System: A Comprehensive Review. Journal of Clinical Medicine. 2024;13(11)doi:10.3390/jcm13113223

46. Arnaud C, Bochaton T, Pépin J-L, Belaidi E. Obstructive sleep apnoea and cardiovascular consequences: Pathophysiological mechanisms. Archives of Cardiovascular Diseases. 2020;113(5):350–358. doi:10.1016/j.acvd.2020.01.003

47. Kario K, Hettrick DA, Prejbisz A, Januszewicz A. Obstructive Sleep Apnea–Induced Neurogenic Nocturnal Hypertension. Hypertension. 2021;77(4):1047–1060. doi:10.1161/hypertensionaha.120.16378

48. Tasali E, Pamidi S, Covassin N, Somers VK. Obstructive Sleep Apnea and Cardiometabolic Disease: Obesity, Hypertension, and Diabetes. Circulation Research. 2025;137(5):764–787. doi:10.1161/circresaha.125.325676

49. Kelmanson IA. Increased P-wave dispersion in patients with obstructive sleep apnea syndrome: a meta-analysis. Sleep and Breathing. 2022;27(1):291–301. doi:10.1007/s11325-022-02630-1

50. Wu C, Huang J, Huang M, et al. Association of electrocardiogram features with risk of obstructed sleep apnea: a population-based cohort study. Sleep and Breathing. 2025;29(1)doi:10.1007/s11325-025-03266-7

51. Qamar U, Jang J-H, Kim TY, Lim H-S, Yoon D. Unsupervised feature learning for electrocardiogram data using the convolutional variational autoencoder. Plos One. 2021;16(12)doi:10.1371/journal.pone.0260612

52. Dykiert IA, Kraik K, Jurczenko L, Gać P, Poręba R, Poręba M. The Effect of Obesity on Repolarization and Other ECG Parameters. Journal of Clinical Medicine. 2024;13(12)doi:10.3390/jcm13123587

53. Cuspidi C, Tadic M, Sala C, Gherbesi E, Grassi G, Mancia G. Obstructive sleep apnoea syndrome and left ventricular hypertrophy: a meta-analysis of echocardiographic studies. Journal of Hypertension. 2020;38(9):1640–1649. doi:10.1097/hjh.0000000000002435

54. Lu M, Wang Z, Zhan X, Wei Y. Obstructive sleep apnea increases the risk of cardiovascular damage: a systematic review and meta-analysis of imaging studies. Systematic Reviews. 2021;10(1)doi:10.1186/s13643-021-01759-6

55. Cloward TV, Walker JM, Farney RJ, Anderson JL. Left Ventricular Hypertrophy Is a Common Echocardiographic Abnormality in Severe Obstructive Sleep Apnea and Reverses With Nasal Continuous Positive Airway Pressure*. Chest. 2003;124(2):594–601. doi:10.1378/chest.124.2.594

56. Garg H, Garg P. Sleep testing in India: Are we missing the opportunity?—A commentary on the need for standardisation and innovation. Lung India. 2026;43(2):113–115. doi:10.4103/lungindia.lungindia_682_25

57. Kim D, Han JY, Jung H, et al. AI-Enhanced Smartwatch AHI Estimation and AI-Scored Polysomnography for Obstructive Sleep Apnea: Real-World Validation. Nature and Science of Sleep. 2025;Volume 17:2297–2307. doi:10.2147/nss.S540460

